# Stable *Plasmodium falciparum* merozoite surface protein-1 allelic diversity despite decreasing parasitemia in children with multiple malaria infections

**DOI:** 10.1101/2024.05.30.24308208

**Authors:** Reuben M. Yaa, Kelvin M. Kimenyi, Palasciano H. Antonio, George Obiero, Lynette Isabella Ochola-Oyier

## Abstract

Individuals experiencing recurrent malaria infections encounter a variety of alleles with each new infection. This ongoing allelic diversity influences the development of naturally acquired immunity and it can inform vaccine efficacy. To investigate the diversity and infection variability, *Plasmodium falciparum* merozoite surface protein 1 (*Pf*msp1), a crucial protein for parasite invasion and immune response, was assessed in parasites isolated from children with repeated malaria episodes. A total of eleven microhaplotypes were observed across all malaria episodes. There were 4 prevalent microhaplotypes, E-KSNG-L, Q-KSNG-L, Q-KSNG-F, and Q-KNNG-L, in the population. Conversely, microhaplotypes such as E-KSSR-L, E-KNNG-L, E-KSSG-L, E-TSSR-L (3D7), Q-TSSR-L, E-TSSG-L, and E-KSNG-F were rare and maintained at low frequencies. High allelic replacements were observed, however some individuals experienced consecutive re-infections with the same microhaplotype. Notably, *Pf*msp1_19_ allelic diversity as measured by haplotype diversity was stable, while nucleotide diversity decreased over time with decreasing parasitemia. Parasite *Pf*msp1_19_ allelic diversity remained stable over the multiple malaria episodes, despite declining parasitaemia levels and ongoing immune development. In addition, our findings reveal dynamic *Pf*msp1_19_ allelic replacements across parasite infection episodes, with initial immune responses to prevalent variants potentially waning over time, allowing for re-infections. Overall, our findings emphasize that protective immunity to malaria involves complex immune responses targeting multiple alleles that wane over time and thus the development of sterilizing immunity is a challenge.

## Introduction

In an attempt to evade host immunity during infection, *Plasmodium falciparum* parasites regularly replace merozoite antigen epitope conformations and thus have the option to use alternative invasion pathways, disrupting complement activation (Herrera et al. 2015; Awandare et al. 2018; Larsen et al. 2018; Oyong et al. 2018). At the genetic level, the changes may arise from point mutations leading to single nucleotide polymorphisms (SNPs), insertions/deletions of one or more base residues as well as meiotic recombination events of parental alleles, generating newer progeny (Mu et al. 2005; Miles et al. 2016). This is further maintained by the phenomenon of balancing selection. This process stabilizes the polymorphic circulation of immune targeted antigens resulting in multi-allelic circulation of these genes especially in malaria-endemic regions (Ochola-Oyier et al. 2019). Effects of balancing selection have been shown before in merozoite surface protein (MSP) 1 (Parobek et al. 2014), apical membrane antigen-1 (ama1) (Polley and Conway 2001), MSP3 (Polley et al. 2007), erythrocyte binding antigen-175 (EBA-175) (Verra et al. 2006), MSP Duffy binding Ligand-1 and 2 (MSPDBL1 and MSPDBL2) (Tetteh et al. 2009; Ochola et al. 2010), reticulocyte binding homologues-2 (Rh2) (Rayner et al. 2005; Reiling et al. 2010) and Rh5 (Ochola-Oyier et al. 2016).

Merozoite antigen alleles are maintained by frequency-dependent immune selection which shifts allele frequencies over time. This is because the rare alleles are observed less and therefore rise to high frequency (Conway et al. 1992). This pattern of selection supports allele-specific immunity, which has repeatedly been shown to reduce overall vaccine efficacies that are based on low frequency or single alleles in different malaria endemic settings (Takala et al. 2007a; Ogutu et al. 2009; Ouattara et al. 2013). Of interest is *P falciparum* MSP1, which has been advanced over time as a vaccine candidate and it is a suitable marker for genotyping parasite populations in antimalarial efficacy studies and clinical trials (Mwingira et al. 2011). However, allelic replacements are associated with reduced efficacies on vaccine formulations targeting this antigen (Rotman et al. 1999; John et al. 2004a; Chauhan et al. 2010; Wilson et al. 2011).

*Pf*MSP1 is a predominant antigen on the surface of the asexual blood stage of the parasite that plays an imperative role in erythrocyte invasion to cause malaria clinical symptoms. It is synthesized as a large precursor during schizogony and subsequently processed via proteolytic cleavage into 5 fragments of which the smallest is a 19kDa fragment (*Pf*MSP1_19_). This fragment has two epidermal growth factor (EGF) domains, one located at the C-terminal and another at the N-terminal ends. The C-terminal interacts with band 3, the erythrocyte receptor, to facilitate parasite erythrocyte invasion (Cowman and Crabb 2006). Inside the erythrocyte, the parasite multiplies and later egresses into the bloodstream following the rupture of the erythrocyte, a process in which *Pf*MSP1_19_ is also involved (Tan and Blackman 2021). During egress, subtilisin-like (SUB1) parasite serine protease modifies the structure of *Pf*MSP1 to bind spectrin, a component of the host erythrocyte cytoskeleton to facilitate egress (Das et al. 2015). Genetic diversity studies of *Pf*MSP1 have highlighted that fewer polymorphisms are located at the 19kDa fragment than the rest of the protein, a total of 6 polymorphic loci (Takala et al. 2007b). Probably, because of its direct proximal interaction with its receptor. The 19kDa fragment is easily accessible to the host immune system as evidenced by merozoite invasion and parasite growth inhibition with antibodies in *in vitro* and mice experiments (Rotman et al. 1999; Cowman and Crabb 2006; Takala et al. 2007b). The fragment elicits both humoral and cell mediated immune responses during exposure to natural infections (Egan et al. 1999; John et al. 2004b), particularly to the polymorphic amino acids at the second EGF-like domain (Egan et al. 1995).

Allelic diversity of *Pf*MSP1 at C-terminal region, has been shown previously in malaria endemic regions such as Kenya, Tanzania and Uganda (Egan et al. 1995; Langhorne et al. 2008; Kariuki et al. 2013; Simpalipan et al. 2014). Similarly, significant epitope diversity through immune assays (Conway et al. 1992) have been demonstrated before in longitudinal studies. Though *Pf*MSP1 allelic diversity and patterns have previously been investigated in different malaria endemic regions, it has not been assessed in recurrent multiple infections in moderate to high malaria transmission regions. To achieve this, allelic replacements and the distribution of C-terminal *Pfmsp1* microhaplotypes were determined over time in multiple infections to describe *P. falciparum* infection diversity. Interrogating *Pfmsp1* microhaplotypes in individual infections over time will shed light on the development of naturally acquired immunity with repeated exposures to infection, while also providing valuable guidance for vaccine design and potential optimal intervals for vaccine dosing.

## Methods

### Study design

We retrieved a total of 426 blood samples from 33 children (comprising 19 males and 14 females) who experienced a minimum of 10 malaria episodes. These children were from a cohort that was originally part of a larger integrated study on natural immunity to malaria established in 2005 (Mwangi et al. 2005), in Junju, Kilifi County, Kenya, conducted under institutional ethical review (SERU 3149) with sampling done from 2008-2013. A blood sample was obtained from every participant upon confirmation of a febrile malaria episode. From the blood samples malaria parasitemia load was estimated using microscopy. In this study, the blood samples were used to evaluate *Pf*MSP1 C-terminal coding region from the parasite isolates.

### DNA extraction, PCR and sequencing

Total genomic DNA from the 426 blood samples were extracted using the QIAamp Blood Mini Kit (Qiagen). The 272bp *Pf*msp1 19kDa coding region was amplified by Polymerase Chain Reaction (PCR) using High Fidelity Taq polymerase (Sigma Aldrich, cat. no:11732641001) with *Pf*msp1_19_-F 5’-CAATGCGTAAAAAAACAATGTCC-3’ and *Pf*msp1_19_-R 5’-TTAGAGGAACTGCAGAAAATACCA-3’ specific primers pairs on cycling conditions as follows: 1 cycle at 94<u>°</u>C for 2 minutes, 9 cycles of 94<u>°</u>C for 30 seconds, 44<u>°</u>C for 30 seconds, 72<u>°</u>C for 2 minutes, 24 cycles of 94<u>°</u>C for 30 seconds, 44<u>°</u>C for 30 seconds, 72<u>°</u>C for 2 minutes + 5 seconds per cycle and a final step of 72<u>°</u>C for 2 minutes. The amplified PCR products were separated by 2% (w/v) agarose gel electrophoresis in a buffer composed of 40mM Tris, 1mM EDTA and 20mM Acetic acid (TAE), pH 8.2, for 40 minutes at 100V. PCR products were visualized using 1% SYBR (v/v) safe stained agarose gels and cleaned using ethanol precipitation. Sequencing templates were prepared using BigDye^TM^ Terminator v3.1 cycle sequencing kit. A volume of 3μl of the purified products was resuspended with, 4μl BigDye^TM^ Terminator 3.1 ready reaction mix, 1μl of 10μM of *Pf*msp1_19_-F primer used during fragment isolation and 3μl of deionized water to a total reaction volume of 10μl in 96 well plate. Cycle sequencing of the amplicons was done using PCR as follows: 96<u>°</u>C for 1 minute, 25 cycles of 96<u>°</u>C for 10 seconds, 50<u>°</u>C with +1<u>°</u>C/second for 5 seconds and 60<u>°</u>C for 4 minutes. Sequencing reactions were purified using ethanol/EDTA precipitation and reactions resuspended in Hi-Di^TM^ formamide. The plates were then analyzed using capillary electrophoresis on ABI 3500XL Genetic Analyzer outsourced from Inqaba Biotechnical Industries (Pty), South Africa. Sequences were assembled, trimmed and edited using Sequencher® 5.3 DNA analysis software (Gene Codes Corporations, Ann Arbo, MI USA) and CLC sequence viewer version 7(QIAGEN). DNA sequence data and corresponding translated protein were aligned to *P. falciparum* 3D7 *msp1* (PF3D7_0930300) msp1 (PF3D7_0930300) reference sequence, ASM276v2, using the MUSCLE alignment algorithm in the MEGA 11 program (Edgar 2004; Tamura et al. 2021). The sequences were deposited in the GenBank NIH genetic sequence database under accession numbers (OQ821998 - OQ822147).

### Data processing and statistical analyses

After standardizing the sequences to the same length (234bp) and excluding short sequences that did not cover the segregating sites in either orientation, sequences were clustered using USEARCH v11 software (Edgar 2010) and Phyclust R package (Tzeng 2005) to identify microhaplotypes. This was followed by determining microhaplotype frequencies. Phyclust applies grouping of microhaplotypes and categorizes those to be retained above a cut-off point which is an optimal balance between the sample size, microhaplotype number and frequencies (Tzeng 2005).

Microhaplotype sequences were extracted as an alignment and transformed to a DNAbin object using ape R package (Paradis et al. 2004). The object was transformed to a hamming distance matrix by measuring pairwise distances of corresponding residues between microhaplotype pairs, while counting differences between them and storing this on a symmetric matrix which was visualized as a heatmap (Pinheiro et al. 2005).

Temporal microhaplotype distribution densities, frequencies, and patterns at the individual and population levels were assessed. A likelihood ratio test was used to determine whether the haplotypes were uniformly distributed in the population (Read and Cressie 2012). More specifically, modeling microhaplotype occurrences using a multinomial distribution, the test evaluated uniform probabilities of infections and compared them to probabilities computed via maximum likelihood estimation, providing strong evidence in favour of the latter. The expected number of microhaplotypes per infection episode was estimated as *p_i_* × *N_t_*, where *p_i_* denotes the maximum likelihood estimate of the probability of seeing haplotype *i* and *N_t_* the total number of observations at episode *t*. Where the expected versus observed differed by a large amount, we tested if the discrepancies were significant using Holm’s method to control for the family-wise error rate (Holm 1977).

In order to characterize the distribution patterns of microhaplotype across the infections, we employed kernel density probability estimates (Węglarczyk 2018), line graphs of microhaplotype frequencies as proportions of the total number of patients and infection variability using the microhaplotype data. The microhaplotype distribution densities and line graphs were compared between prevalent and rare microhaplotypes. A population infection variability analysis was conducted to examine the genetic diversity of the *Pfmsp1* locus using haplotype and nucleotide diversity indices. We assigned microhaplotypes back to the patients to assess patient-microhaplotype distribution and proportions. The time between infections for individuals was determined by calculating time elapsed between successive infections.

To determine parasitemia and microhaplotype dynamics over time, the outcome of *Pfmsp1_19_* PCR on all the samples and genetic diversity of the microhaplotype was correlated using the spearman rank method with malaria parasitemia. To do so, we only used data corresponding to the first 14 episodes, since the amount of data collected beyond this episode was insufficient to appropriately carry out this analysis. In addition, microhaplotypes that did not have respective parasitaemia data were excluded from the analysis. The samples were grouped based on *Pfmsp1_19_* PCR amplification status as either amplicon present or absent. Parasitemia was determined by microscopic examination of blood films of *P. falciparum* parasites by counting the number of parasites/200 white blood cells (Mwangi et al. 2005) (**Supplementary Table 1**). The difference in parasitemia between the two groups was compared using the Wilcoxon rank-sum test. We assessed the correlation of parasitemia and microhaplotype diversity as a measure of infection on time dimension.

## Results

### Microhaplotype classes and associated specific patterns in the population

We genotyped the *Pfmsp1_19_* fragment from blood samples of 33 children (19 males and 14 females) with infections spread between 10 and 24 multiple episodes totaling to 426 infections. On recruitment, the average age of the participants was (5.5 ± SD 1.8 years), and at the end of study was (10.30 ± SD 1.8 years). A total of 64.8% (276/426) of the samples yielded *Pfmsp1_19_* amplicons, of these, a total of 65.2% (180/276) were sequenced and only 54.3% (150/276) generated the full length (234bp) Pfmsp119 C-terminal contigs that were used for the microhaplotype analysis. The median parasitemia was significantly higher among in the samples for which *Pfmsp1_19_* amplicons were generated, 160,000 parasites/µl (interquartile range (IQR) = 245,740) compared to those where no *Pfmsp1_19_* amplicons were generated, 15,800 parasites/µl (IQR = 113,640) (P < 0.0001). Throughout the entire period, we genotyped an average of 5 samples (± SD 2.678) per individual, which yielded a total of 150 sequences. We identified six distinct nucleotide polymorphisms at positions 4990, 5132, 5157, 5159, 5161, and 5206 in relation to *Pfmsp1* reference gene coordinates. The nucleotide substitutions in the polymorphic sites resulted in non-synonymous amino acid substitutions at codons 1644, 1691, 1699, 1700, 1701 and 1716 yielding a total of 11 microhaplotypes (**Figure 1A**), all of which have been reported in previous studies (Roy et al. 2008; Mwingira et al. 2011). Microhaplotypes E-KSNG-L, Q-KSNG-L, Q-KSNG-F and Q-KNNG-L corresponding to FUP-Uganda PA, FVO Wellcome, Thai (T807) and Kenya strains respectively, were the dominant microhaplotypes circulating in the population with proportion frequencies of 36% (54/150), 26% (39/150), 18% (27/150) and 4.7% (7/150), respectively and these microhaplotypes were classified as prevalent. Both E-KSSR-L and E-KNNG-L haplotypes were circulating with frequencies of 4% (6/150), and together with the remaining microhaplotypes, E-KSSG-L, E-TSSR-L (3D7), Q-TSSR-L, E-TSSG-L and E-KSNG-F with low frequencies of <5%, were considered as rare circulating microhaplotypes in the population (**Figure 1B**).

**Figure 1.**
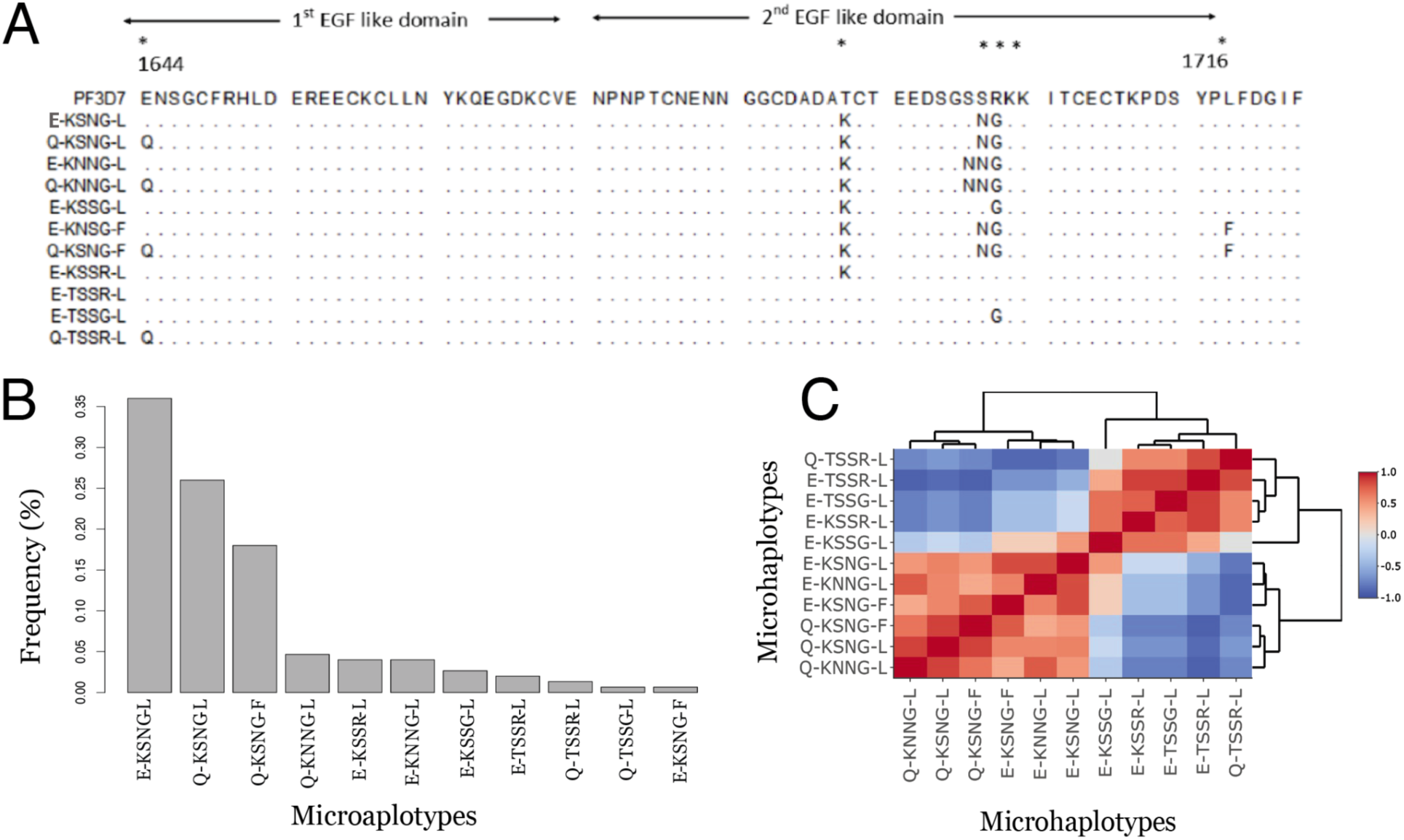
Dynamics of *P. falciparum* msp1_19_ microhaplotypes. (**A**) Amino-acid sequence alignment of 11 identified microhaplotypes. Polymorphic sites are shown with an asterisk (*). The nucleotide positions relative to the start position of the *Pfmsp1* gene are shown below the asterisk. The dots in the alignment indicate the position corresponding to *P. falciparum 3D7* with identical amino acid sequences. The epidermal growth factor (EGF)-like domains 1 and 2 are shown by arrows. The first polymorphism is located in the first EGF-like domain, whereas the second to the fifth polymorphism are located in the second EGF-like domain. (**B**) Microhaplotypes sorted by their abundance in the population. (**C**) Microhaplotypes clustered to groups based on the number of nucleotide differences between haplotypes. The dendrogram on the sides of the heatmap visually represents the relatedness of the microhaplotypes. In this context, the branches indicate distinct clusters formed through hierarchical clustering, highlighting groups of haplotypes with similar characteristics. Scale=Pearson correlation coefficients.

At least 7 of the 33 patients exhibited infections with several different microhaplotypes ranging from 7 to 10 over the entire infection period (**Figure 2A**, **Supplementary Figure 1A**). All patients were infected with at least one or more of the predominant microhaplotypes. Specifically, 72.7% (24/33) were infected with the E-KSNG-L predominant microhaplotype. This microhaplotype ranged from 22.2%-75% per patient in relation to all genotyped re-infections per individual (**Supplementary Figure 1B**).

**Figure 2.**
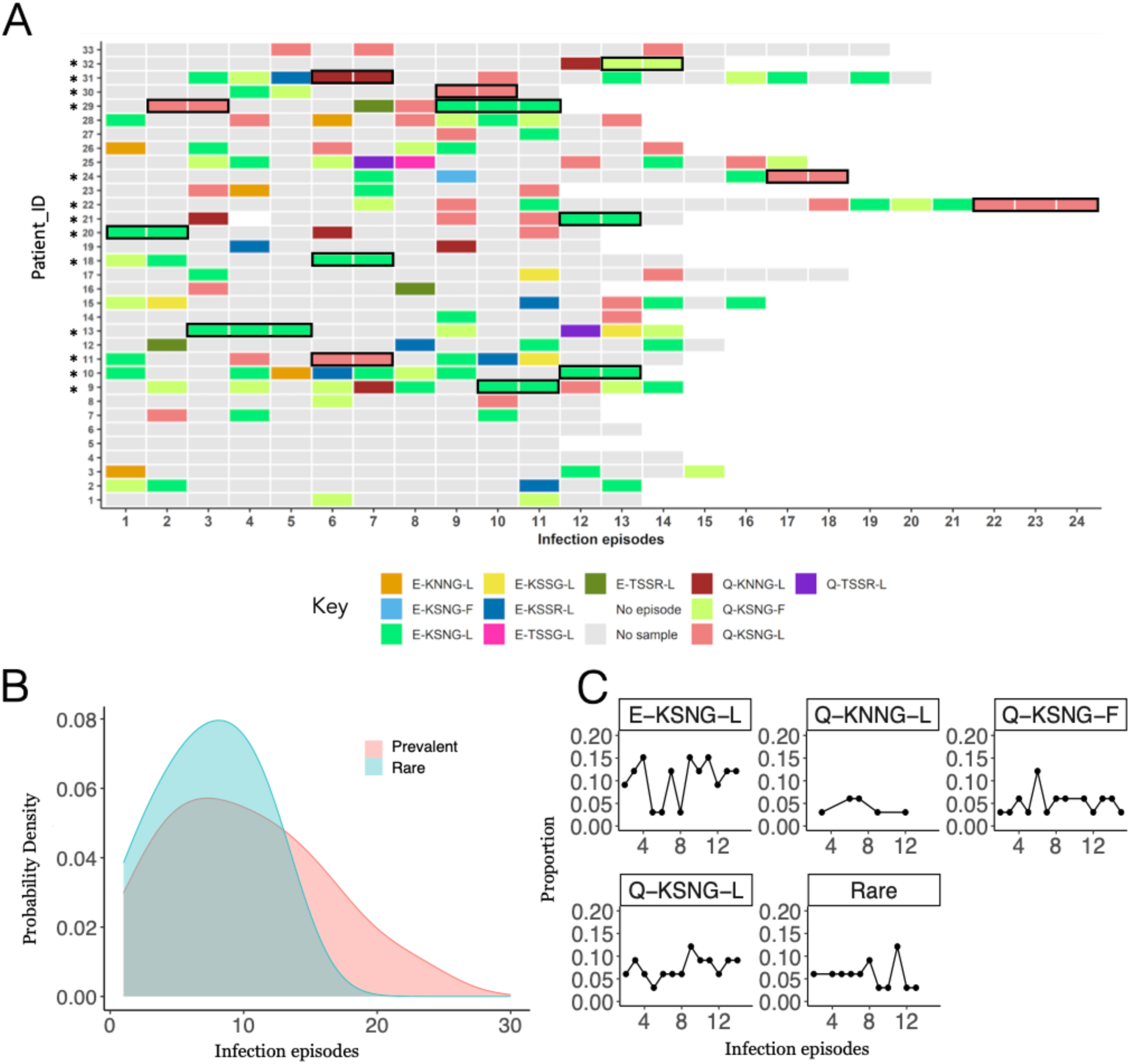
Microhaplotype patterns across the infections(**A**). The distribution of microhaplotypes in each patient across the malaria episodes. No sample - Samples not retrieved from the biobank, samples not genotyped or sequenced. No episode - No malaria episode was reported. Patient_IDs with asterisks represent cases that had 2-3 similar consecutive microhaplotypes outlined in black. (**B**) Density of prevalent (≥ 5% frequency) and rare (<5% frequency) microhaplotypes across the infection episodes based on Figure 1B. (**C**) Line graph of proportions of prevalent microhaplotypes (E-KSNG-L, Q-KSNG-L and Q-KSNG-F), and rare microhaplotypes across the first 15 infection episodes. Proportions are expressed from the total children genotyped at each infection episode.

The microhaplotype hamming distance matrix classified the microhaplotypes into 3 groups based on Pearson correlation measures (**Figure 1C**). The first larger group was composed of E-KSNG-L, E-KNNG-L, Q-KSNG-L, Q-KSNG-F, E-KSNG-F and Q-KNNG-L and these were the prevalent microhaplotypes. The second group was made up of a single microhaplotype, E-KSSG-L, and the third group included E-KSSR-L, E-TSSG-L, E-TSST-L and Q-TSSR-L microhaplotypes which were the rare circulating microhaplotypes (**Figure 1C)**.

### Microhaplotype dynamics over time and across infections

We followed the pattern of the microhaplotypes across infections to investigate the allelic replacements of the C-terminal of *Pf*msp1_19_ over the course of multiple infections. At an individual level, there were notably high random allelic replacements between re-infections. At least 39.4% (13/33) of the patients were consecutively re-infected with the same microhaplotype of the prevalent alleles either 2 to 3 times across the infection (**Figure 2A**). With the exception of one case (1/14) the consecutive re-infections occurred within a one-year timeframe **(Figure 2A**, **3B**). Remarkably, all individuals experiencing consecutive re-infections with the same microhaplotype apart from one (1/13), showed no recurrence of those specific microhaplotypes in subsequent parasite infections (**Figure 2A**). The average interval between infections for the entire period was 5.0 months, ranging from 1 and half weeks to around 25 months (**Figure 3A**).

**Figure 3:**
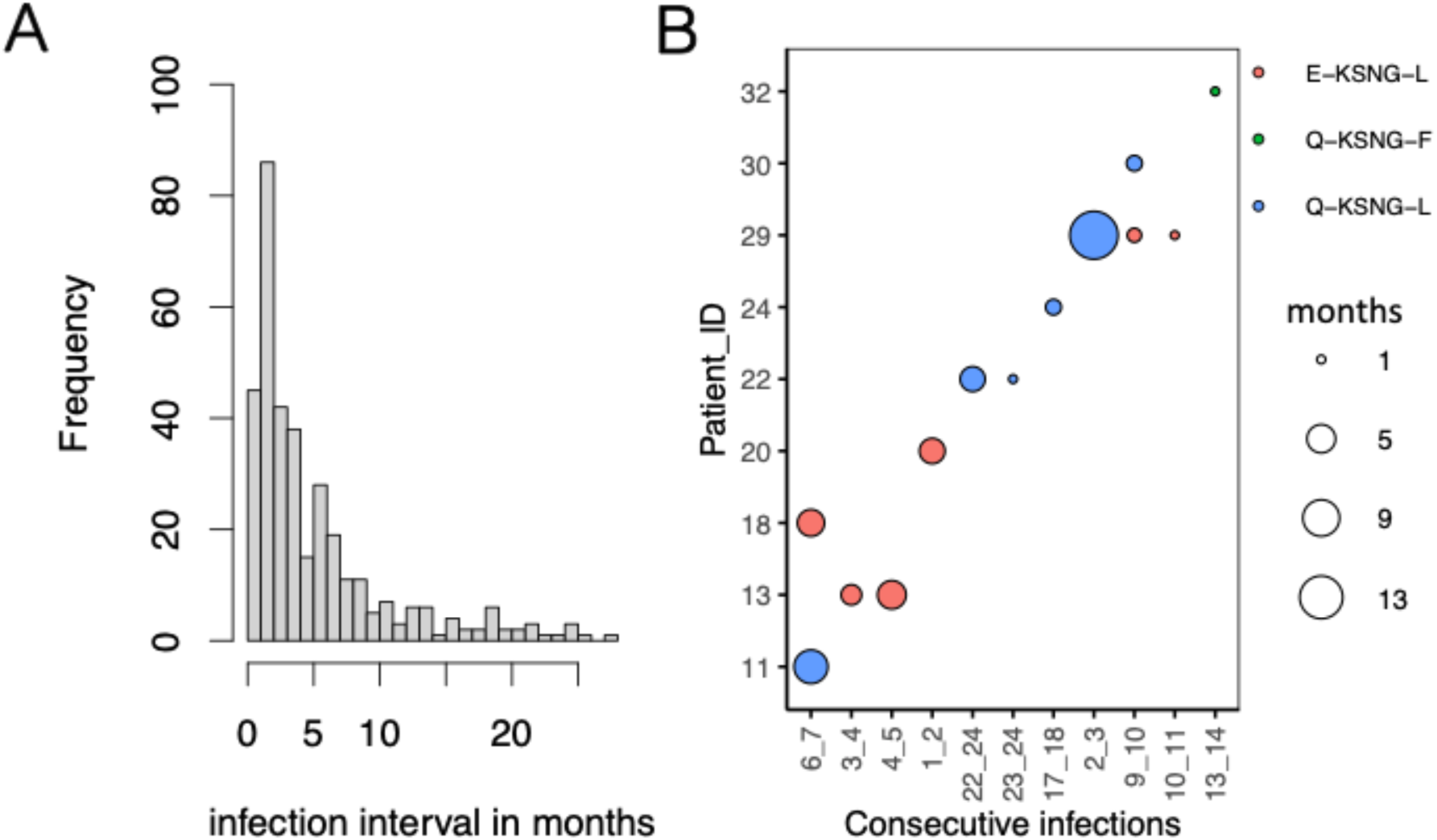
Distribution of infection intervals. **(A**) Frequency histogram of the distribution of the time interval between infections in months for all infections with genotype data. **(B)** Interval in months for individuals with consecutive infections of the same microhaplotype. The size of the circles depicts the number of months between infections with the same microhaplotype depicted by the color of the circle.

At a population level there was a peak in microhaplotype density before the 10th infection episode, with the rare alleles peaking slightly later just after episode 10 (**Figure 2B**). We checked the pattern of the four most frequent microhaplotypes and rare microhaplotypes across the multiple infections. Of these, microhaplotypes E-KSNG-L was observed throughout the infections, Q-KSNG-L, Q-KNNG-L and Q-KSNG-F peaked once before episode 10. The rare microhaplotypes peaked after episode 8 (**Figure 2B, C**). We noted that the occurrences of the prevalent microhaplotypes was not influenced by chance as the observed frequencies matched the calculated expected frequencies (**Supplementary Figure 2)**.

### Correlation of parasitemia, genotyping and microhaplotypes

The change in parasitemia levels was examined over time and correlated with microhaplotype and nucleotide diversities across the infection episodes. During the early and middle stage infection episodes (<8 episodes) parasitemia levels were notably high. However, as the infections progressed from towards the later (>8) episodes, parasitemia levels exhibited a decreasing trend. Genetic diversity of the locus remained stable over the course of infection episodes (**Figure 4**). These trends were found to be positively correlated with parasitaemia, with a strong positive correlation coefficient of 0.7 for nucleotide diversity and but not for microhaplotype diversity, 0.37.

**Figure 4.**
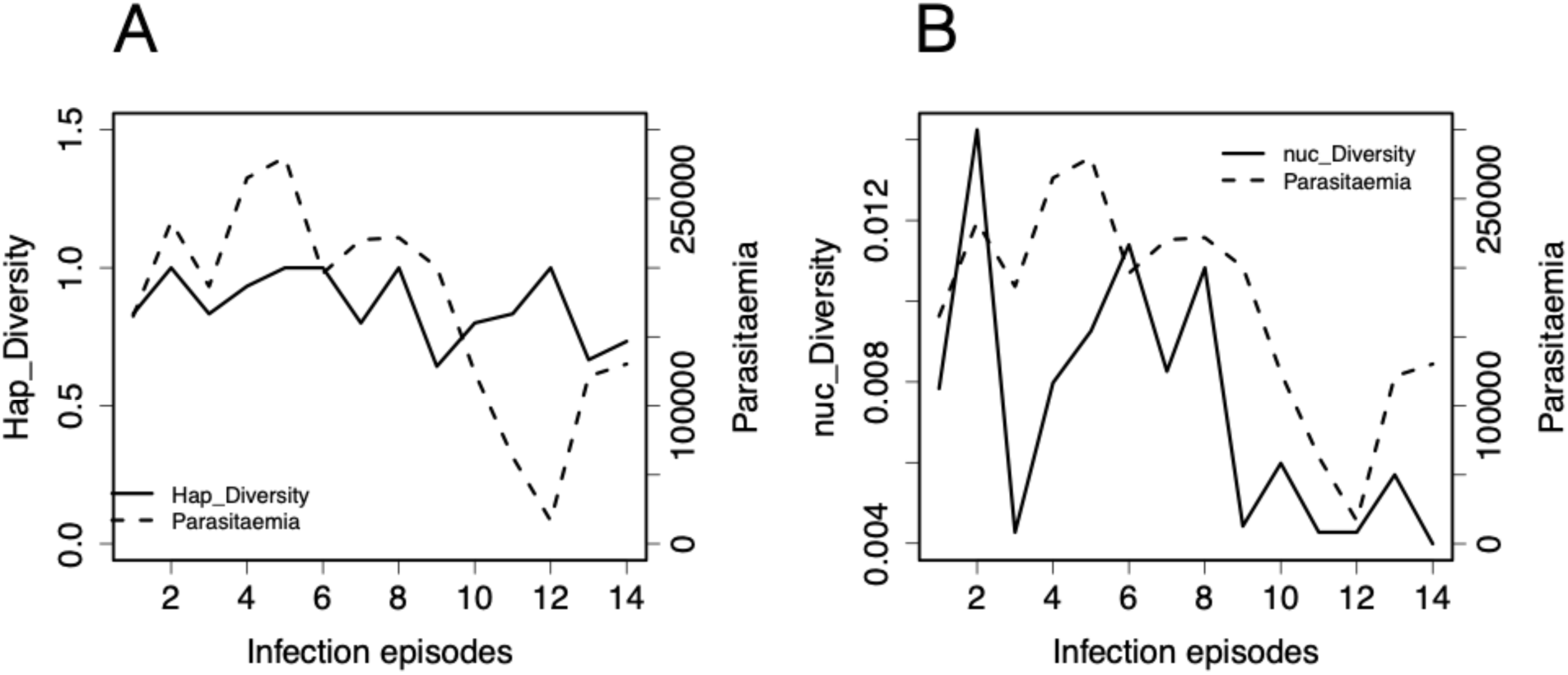
Parasitaemia correlations with measure of genetic diversity across infection episodes. (**A**) Haplotype diversity (hap_diversity) fluctuates within a small range (between 0.6 and 1) across infection episodes as parasitaemia (parasites/µl) reduces, the correlation between haplotype diversity and parasitemia was low, 0.37. (**B**) Nucleotide diversity (nuc_diversity) reduces concurrently across infection episodes with parasitaemia with a correlation of 0.7.

## Discussion

Given the dynamic interplay between host immune responses which continuously shape merozoite antigen diversity (Ferreira Marcelo U. et al. 2004; Early et al. 2018; Ochola-Oyier et al. 2019; Naung et al. 2022) by shifting allele frequencies and favoring the presence of rare alleles, we leveraged the diversity of *Pfmsp1* in parasite isolates from children with multiple malaria infections (a proxy of developing immunity) as a window into quantifying the perturbation on merozoite antigen allele diversity. Our results revealed preserved genetic diversity in *Pfmsp1*, even as parasitaemia levels decreased over multiple malaria infections in an individual. This suggests that long-term immune pressure does not alter the overall allele diversity. Similar patterns especially for the msp1-19 microhaplotypes have been observed across Sub-Saharan Africa including in the Coast of Kenya, Western Kenya, Republic of Congo, Uganda, Tanzania, Mali and Burkina Faso (Mwingira et al. 2011; Kang et al. 2012; Kariuki et al. 2013; Ochola-Oyier et al. 2019; Baina et al. 2023) suggesting that in moderate to high transmission, *P. falciparum* populations maintain a complex infection pattern that supports out-breeding while preserving survival genetic diversity.

It is expected that following multiple exposure to a single allele, immunity develops and reduces its frequency in subsequent infections. However, repeat infections with the prevalent alleles circulating in the population was common, with over a third of the children showing consecutive infections with the same allele and only 6 children not showing a repeat infection with the same allele. The limitation in these 6 children is the low number of genotyped samples. Importantly, these findings highlight the complexity of the parasite’s genetic diversity that needs to be determined in the light of other polymorphic antigens. Though the *Pfmsp1* alleles are the same in repeat infections, this region of the antigen is limited in diversity to demonstrate distinct allelic changes. Other genetically diverse antigens may show differences between each infection such as *ama1* or block 2 of *msp1* and block 3 of *msp2*. What was unique in our data was the reduction in parasitemia in later infections, which did not lead to a reduction in haplotype diversity. Though it is clear that following several malaria infections, the host can control parasitemia due to the development of immunity (usually antibody-mediated) (Pohl and Cockburn 2022) with age; there appears to be a lapse in the allele-specific immunity memory, such that over time antibody levels targeted to a particular allele are low or wane over a short period (about <1 year) when re-infections occur with the same allele. It is surprising that with continued exposure and increasing age re-infections with the same alleles occur, while it is expected that exposure to several malaria infections should increase the antibody repertoire to the several antigens and polymorphic combinations presented over time. However, what is evident is the difference between anti-parasite immunity that develops to control parasitemia levels following several infections, and increasing age, in an individual and the immunity that develops to clear allelically different epitopes of the same antigens. The control of parasitemia following several malaria infections is similar to previous findings in Uganda that observed lower parasite densities with increasing age and in high malaria transmission areas (Rodriguez-Barraquer et al. 2018). Thus, emphasizing the difference in the immunity that controls parasitemia and that which could lead to sterile immunity. This latter process is not achieved for malaria, based on this data where repeated exposure by the same prevalent allele occurs that is not completely cleared, potentially due to an ineffective immune response that is not protective, akin to the original antigenic sin hypothesis (Good et al. 1993). There may be some cross-reactivity in responses between alleles, however this too is likely to be limiting since re-infections occur with the prevalent alleles circulating in the population. The re-infections with the same allele allows the maintenance of their high prevalence in the population and thus the genetic diversity of the infections is unaltered. The high haplotype diversity is sustained while the nucleotide diversity in contrast dropped with the parasitemia levels. The nucleotide diversity is likely to reduce as re-infections occur with the same allele, the average nucleotide differences between sequences across the population will thus reduce. However, it will not alter the overall haplotype diversity the probability that two randomly sampled alleles are different in the population. Furthermore, this population is unique in the sense that these are children who are uncharacteristically infected several times over 5 years with malaria and who have been shown to have a modified immune system high immune activation and inflammation, TNF-α, IL-6, IL-10 and cell populations such as γδ T cells were significantly higher in children with >8 malaria infections compared to the <5 infections group (Bediako et al. 2019). This skewed cytokine profile may act in a way that the inflammatory immune response to some extent clears parasites controlling parasitemia, however the IL-10 modulates the inflammatory response limiting inflammation and related immunopathology, but it may interfere with robust protective allele-specific immune responses.

Of additional interest, was the time between infections that was on average 5 months for the recurrent allelic infections, suggesting that immunity then wanes after 5 months and reinfections then occur in this group of children who experience multiple malaria infections. This is likely to be a primer for the design of multi-allelic blood stage vaccine combinations, as potentially giving an individual a cocktail of alleles or alternating between alleles in follow up vaccine doses five or six months apart.

It is worth noting that our observations would have been more robust if complemented with other invasion genes with higher levels of polymorphisms to support interpretability of our findings. In addition, without immunological functional validation experiments, uncertainty persists regarding whether the absence of consecutive same variants in late infections among certain individuals, and distribution densities in later infections are a result of allelic-specific or antigenically dissimilar variant-transcending immune responses. Further experiments in this direction are needed to clarify this observation.

## Conclusion

Parasite *Pfmsp1_19_* allelic diversity remains stable over the multiple malaria episodes despite declining parasitaemia levels over time due to population-wide immune pressure. This suggests that immune responses targeting Pfmsp1 may not be the primary factor driving the observed reduction in parasitaemia, underscoring the multifactorial nature of protective immune responses against malaria. In addition, our findings reveal a high *Pfmsp1_19_* microhaplotype replacement across multiple infections. While shifts in alleles between infections indicate initial immune responses to variants, these responses are not consistently maintained over time, allowing for re-infections with the same variant. In conclusion, the study highlights that immunity to malaria is not achieved through complete sterilizing immunity but rather involves a complex interplay of immune responses targeting multiple alleles from multiple antigens.

## Data Availability

All data produced in the present work are contained in the manuscript and at GenBank NIH genetic sequence
database under the accession numbers:(OQ821998 - OQ822147)

https://www.ncbi.nlm.nih.gov/nuccore/

## Data and code availability

### Data accessibility

The DNA sequences for the C-terminal of *Pf*msp1*_19_* were deposited in GenBank NIH genetic sequence database and are available under the accession numbers: (OQ821998 - OQ822147) (**Supplementary Table 1**).

### Code

Analysis and code for this study is available in the dedicated GitHub repository: https://github.com/mangiruben/pfMSP1-Malaria

## Declarations

### Author contributions

RMY performed all experiments, performed PCR and library preparation, data analysis and wrote the manuscript. KMK did analysis and wrote the manuscript. PHA did analysis and wrote the manuscript. GO oversaw the study and contributed to the manuscript. LIO-O conceived, designed, and oversaw the study and wrote the manuscript. All authors read and reviewed the manuscript.

## Acknowledgements

We thank Ann Owiti and Edwin Rono at Centre for Biotechnology and Bioinformatics (CEBIB), University of Nairobi for overseeing PCR, library generation and sequencing. The Biosciences department at KEMRI-Wellcome Trust Research Programme, Kilfi for the DNA samples from Junju cohort. We thank the previous Directors of CEBIB and the Director of the Kenya Medical Research Institute for permission to publish this article.

## Supplementary Materials

### Supplementary Tables

Table S1. Summary of study participants and blood samples

Supplementary Table 1(excel sheet)

### Supplementary Figures

**Supplementary Figure 1:**
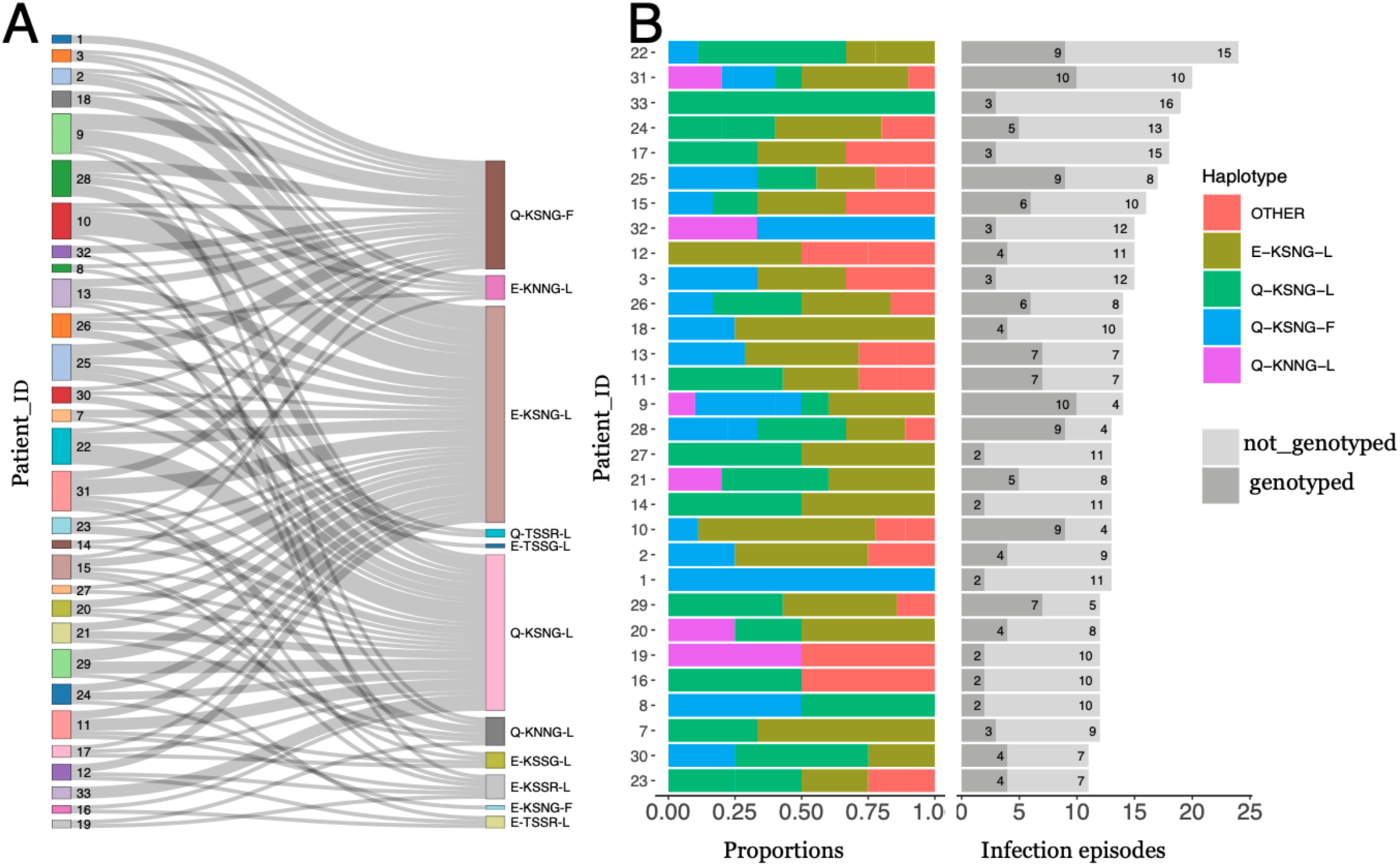
Within host haplotype dynamics. (**A**) Haplotypes assigned back to the patient. Sankey plot showing the assignment of microhaplotypes each patient (Patient_ID) encountered across the multiple episodes. (**B**) The proportion of prevalent haplotypes (E-KSNG-L, Q-KSNG-L, Q-KSNG-F and Q-KNNG-L) in relation to all the rare variant microhaplotypes (OTHER) per patient (Patient_ID). The grey bar panel on the right shows the number of episodes of genotyped and not genotyped per patient.

**Supplementary Figure 2:**
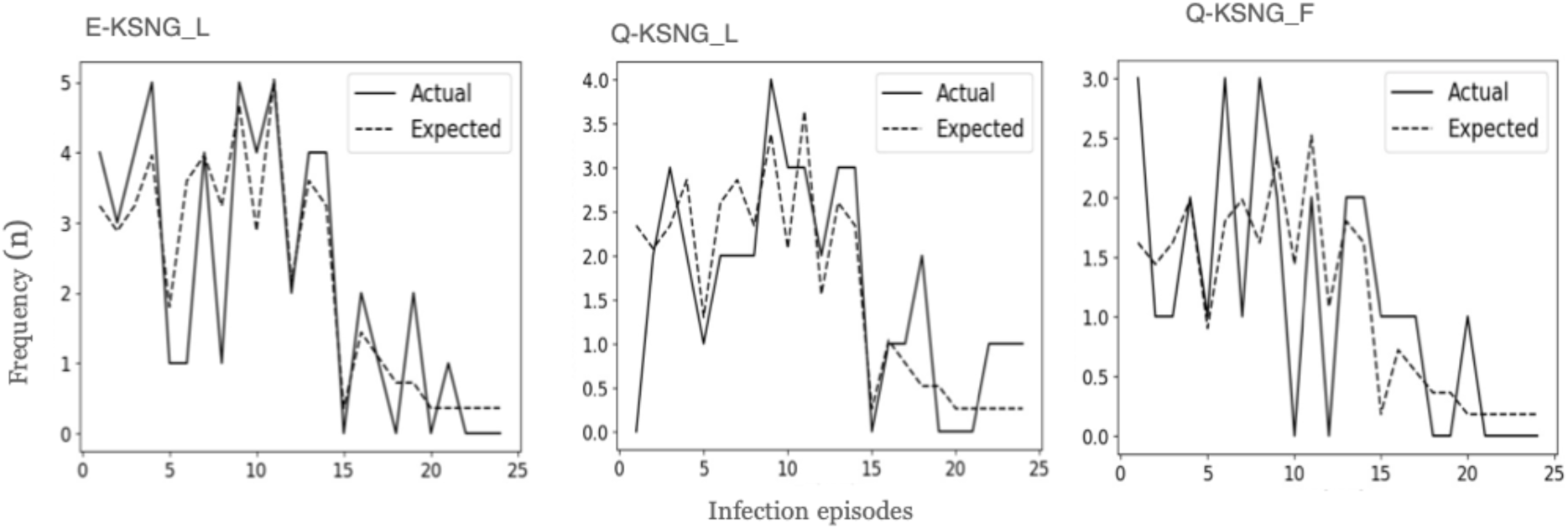
Difference between actual and expected microhaplotype frequencies across infections. The expected number of occurrences was derived using probabilities computed from maximum likelihood estimates.

